# DNA hypohydroxymethylation in pediatric central nervous system tumors is associated with CTCF binding sites and reduced survival

**DOI:** 10.1101/2020.06.20.20136184

**Authors:** Nasim Azizgolshani, Curtis L. Petersen, Youdinghuan Chen, Lucas A. Salas, Laurent Perreard, Lananh N. Nguyen, Brock C. Christensen

## Abstract

Nucleotide-specific 5-hydroxymethylcytosine (5hmC) remains understudied in pediatric central nervous system tumors. We measured genome-scale 5hmC in glioma, ependymoma, and embryonal tumors from children, as well as control pediatric brain tissues using oxidative and bisulfite treatments. Tumor 5hmC localized to regulatory elements crucial to cell identity, including transcription factor binding sites and super-enhancers. A linear model tested the CpG-specific differences in 5hmC between tumor and non-tumor samples, as well as between tumor subtypes. Compared to non-tumor samples, tumors were hypohydroxymethylated across the epigenome. Differentially hydroxymethylated loci among tumor subtypes tended to be hypermethylated and disproportionally found in CTCF binding sites and genes related to posttranscriptional RNA regulation, such as *DICER1*. Model-based clustering results indicated that patients with low 5hmC patterns have poorer overall survival and increased risk of recurrence. These results have implications for emerging molecular neuropathology classification approaches and epigenetic therapeutic strategies in childhood brain tumors.

## Introduction

Central nervous system malignancies are the most common pediatric solid tumor type in North America^1^. They are the leading cause of death from disease in childhood and are the greatest source of cancer-derived morbidity in the US, due to treatment side effects and resistance.^2^ Pathologists commonly classify these tumors into glioma, ependymoma, and embryonal types.^3^ Within these broad groups, there is substantial variability in histopathology and patient prognosis. For instance, gliomas encompass diagnoses such as glioblastoma, a high-grade cancer with low survival requiring chemotherapy, radiation, and surgery, while pilocytic astrocytoma, a benign entity, is cured by surgery alone. Due to the variation in underlying disease mechanism and impact on prognosis, risk stratification and classification are critical to treatment planning and goal setting.

Several studies have paved the way in using molecular markers to classify pediatric central nervous system (CNS) tumors. Through gene sequencing, diagnoses have been subdivided. *WNT1* and *SHH* mutations have been identified as subtype-specific genetic alterations in group 1 and 2 medulloblastomas with subsequent treatment and prognostic implications.^4,5^ Likewise, ependymomas have been split into multiple subgroups based on *C11orf95-RELA* and *Yap1* fusions,^6^ while *BRAF-KIAA1549* fusion has been used to characterize pediatric low-grade gliomas.^7^ These molecular markers have recently been incorporated into the WHO CNS tumor classification guidelines.^3^ The same gene pathways that are implicated in tumorigenesis, such as *SHH, WNT1, NOTCH1*, and *PTEN*, play a critical role in normal neurogenesis and development.^8^ However, while genomic approaches have provided ways to better characterize pediatric CNS tumors, many lack obvious genetic drivers, and these genetic findings are not always associated with prognosis.

Epigenetic profiling to characterize CNS tumors in both research and clinical settings is increasingly applied. DNA methylation, the addition of a methyl group to cytosine, 5-methylcytosine (5mC), is one of the most common and best-studied epigenetic modifications. Alterations in the methylation status of specific cytosines have been used to classify CNS tumors in both adults and children to great effect —identifying changes that transcend somatic genetic alterations.^9–12^ Tumor DNA methylation signatures have been used not only for diagnostic purposes but to stratify subgroups with a worse prognosis.^13,14^ While genome-scale 5mC profiles have been well described and used clinically in CNS tumors; there is very limited nucleotide-specific data for the other major cytosine modification in the brain, 5-hydroxymethylcytosine (5hmC). In mammals, TET proteins oxidize 5mC to 5hmC, which can be further degraded to 5-formylcytosine (5fC) and 5-carboxylcytosine (5caC) leading to methylation loss.^15,16^ Though initially thought to be an intermediate, 5hmC has since been shown to play a distinct role in regulating gene transcription and increasing chromatin accessibility.^17–19^ Unlike methylcytosine, 5hmC concentrates in gene bodies and enhancers and is associated with increased gene expression.^20–22^ 5hmC tends to be depleted in promoter CpG islands and is instead observed in the flanking CpG shores and shelves.^21,23^

5hmC modifications are most abundant in the brain, with levels tenfold higher than other tissues, though varying among regions of the brain.^24^ In the cerebellum, 5hmC accounts for up to 42% of cytosine modifications.^25^ Appropriate patterning of 5hmC is likely critical to normal neurodevelopment, and altered 5hmC has been associated with developmental neuropathologies such as Rett syndrome, autism, and schizophrenia.^26–28^ 5-hydroxymethylcytosine has been studied in adult brain tumors, such as glioblastoma, and has been shown to correlate with survival.^20,29,30^ However, genome-wide nucleotide level studies in pediatric central nervous system tumors are lacking. Changes in 5-hydroxymethylcytosine in tumors have been consistent across tissue types, with depletion of 5hmC in gene bodies and regulatory regions such as enhancers and transcription factor binding sites.^31–35^ Loss of 5hmC tends to coincide with commonly mutated genes in cancers and is tissue specific.^17,20,36,37^ 5hmC appears to play a unique role in cancer, such as localizing to sites of DNA damage, and promoters with 5hmC have been shown to be resistant to the hypermethylation that is common in cancers.^35,38^ To date, the scientific community has yet to adequately measure and understand the role of 5hmC in pediatric CNS tumors.

Current methods of measuring nucleotide level 5mC – including those for CNS tumor classification – rely on bisulfite conversion techniques which cannot distinguish between 5hmC and 5mC.^39^ Most studies of 5hmC are based on immunohistochemistry testing and only provide a genome-level summary measure of 5hmC, without chromosome, gene-region, or nucleotide specific information.^40,41^ Due to the abundance of 5hmC in the brain and observed alterations to it in tumorigenesis, we aimed to examine 5hmC in pediatric CNS tumors. We measured 5-methylcytosine and 5-hydroxymethylcytosine epigenome-wide at nucleotide resolution in pediatric glioma, ependymoma, and embryonal tumors as well as non-tumor tissue using tandem bisulfite and oxidative bisulfite sequencing followed by hybridization to the Illumina Methylation EPIC Array that interrogates over 860,000 CpG loci.

## Results

### Overview of Study Population

All cases included in this study (n=27) received treatment at the Children’s Hospital at Dartmouth Hitchcock and the Norris Cotton Cancer Center between 1993 and 2009. The mean age of cases was 9.5 years, and the mean age of subjects providing non-tumor samples was 6 years. Tumor samples included thirteen gliomas, eight ependymomas, and six embryonal tumors. Additional details of subject demographic and tumor characteristics are provided in **Table 1**, and tumor diagnostic information is available in **Supplementary Table 1**. Our cohort included both common and rare entities, including dysplastic gangliocytoma and spinal myxopapillary ependymoma. All samples were *H3F3A* (*K27* and *G34*) and *TERT* promoter (C228T and C250T) wild-type, as determined by Sanger sequencing **(Supplementary Table 2)**.

**Table 1.** Patient Demographics and Tumor Characteristics

|  | Tumor (n=27) | Non-Tumor (n=3) |
| --- | --- | --- |
| Age, mean (SD) | 9.5 (5.5) | 6 (5.6) |
| Range | 1-18 | 0 - 11 |
| Sex n (%) |  |  |
| Female | 12 (44) | 1 (33) |
| Male | 15 (56) | 2 (67) |
| Tumor Type, n (%) |  |  |
| Embryonal | 6 (22) |  |
| Glioma | 13 (48) |  |
| Ependymoma | 8 (30) |  |
| Grade n (%) |  |  |
| I | 12 (44) |  |
| II | 5 (19) |  |
| III | 2 (7) |  |
| IV | 8 (30) |  |
| Sample Location n (%) |  |  |
| Supratentorial | 9 (33) | 3 (100) |
| Subtentorial | 17 (63) |  |
| Spinal | 1 (4) |  |
| Median Follow Up (yrs) | 14 |  |
| Mean Time to Recurrence (yrs) | 1.5 |  |

### Pediatric CNS Tumors Are Globally Depleted of 5hmC

To measure nucleotide-specific 5hmC and 5mC levels, we applied tandem bisulfite and oxidative bisulfite treatment to DNA from 27 fresh frozen pediatric CNS tumor samples and three non-tumor control tissues. Treated DNA was hybridized to the Illumina Human Methylation EPIC array. Following processing, CpG probes associated with SNPs and sex chromosomes were removed, and 743,461 CpG sites remained in the dataset.

To first compare our findings with prior work where 5hmC measures only provide a single epigenome-wide summary value, we calculated median 5hmC levels using the 743,461 CpG sites in our dataset. Median 5hmC levels were significantly lower in tumors (gliomas 1.75%, ependymomas 1.76%, and embryonal tumors 1.22%), compared to non-tumor tissues (4.81%) (**Figure 1a**). Median levels of tumor 5mC (glioma 62.5%, ependymoma 62.4%, embryonal 57.7%, non-tumor 60.3%), did not differ from controls (**Figure 1b**).

**Figure 1:**
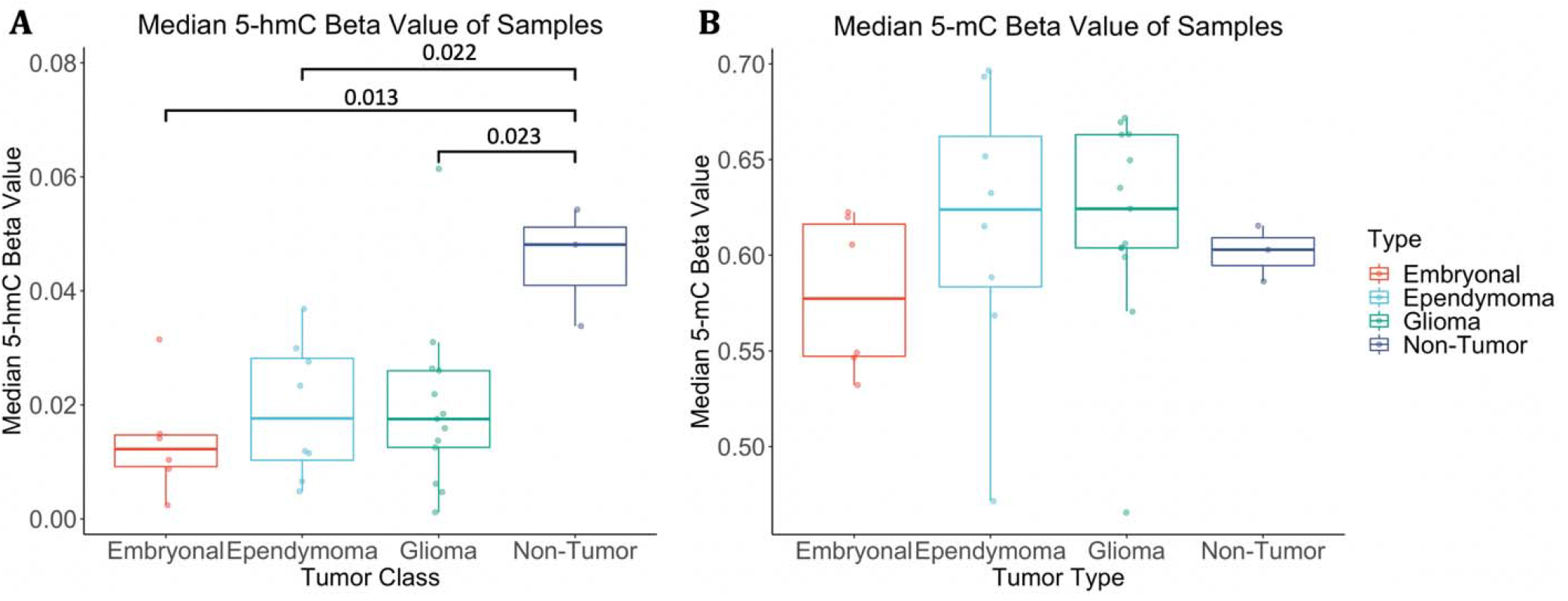
Cytosine modifications in CNS tumors as compared to non-tumor brain tissue. **a)** Consistent with studies in other tumor types, 5-hydroxymethylation levels are depleted compared to non-tumor samples; this depletion holds across tumor types. Student’s t-test comparing median values was performed and differences met statistical significance. **b)** Total methylation levels do not differ significantly between tumor and non-tumor samples.

Cumulative density plots for median 5hmC and 5mC across all tumor samples demonstrated very low 5hmC levels at the majority of CpG sites in contrast to 5mC levels, which were more evenly distributed (**Figure 2a**). We observed a negative correlation between 5hmC and 5mC for approximately 80% of CpGs in tumor samples (Pearson and Spearman’s correlation) (**Figure 2b**). CpG specific median beta values were calculated across all tumor samples and ordered by increasing value to identify the distribution of 5hmC level and loci with appreciable 5-hydroxymethylation (**Figure 2c**).

**Figure 2.**
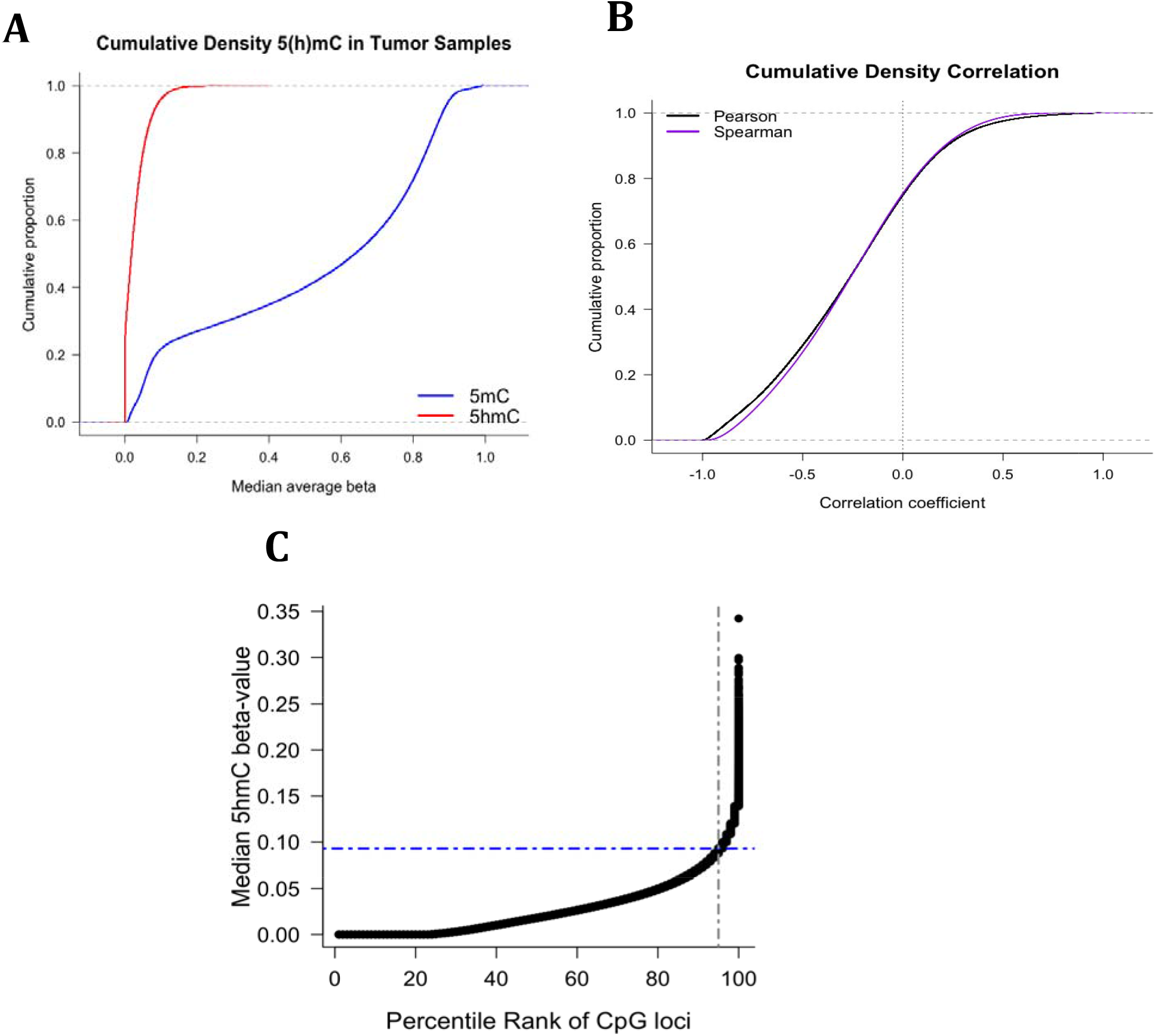
**a)** Empirical cumulative distribution of median 5-hydroxymethylcytosine and 5-methylcytosine across 27 primary pediatric central nervous system tumors. **b)** Cumulative proportions of Spearman and Pearson correlation coefficients calculated for each CpG across all tumors. **c)** Ordered distribution of CpG-specific median 5hmC values across the EPIC array in tumor samples. The x-axis represents percentile rank of CpG by median beta value. We isolated CpGs with medians greater than the 95^th^ percentile and called these high 5hmC CpGs. This corresponded to 37, 173 loci with a median beta value greater than 0.09. We based subsequent analyses on these high 5hmC CpGs.

Levels of 5hmC and 5mC differ between tumors and controls depending on the distance from the transcription start site (TSS). Immediately surrounding the TSS, there was no difference between tumors and non-tumor tissues, and levels of both cytosine modifications drop to near zero. At approximately 300 base pairs in the 5’ direction and 700 base pairs at 3’, tumors demonstrate hypohydroxymethylation and hypermethylation (**Figure 3**). Compared with non-tumor control tissue we observed consistently lower levels of 5hmC within each tumor subtype (**Supplementary Figure 1a**). Despite lower median 5hmC near TSSs, a substantial proportion of high 5hmC CpGs are found within 5kb of the TSS, with 14% of loci within 2000 base pairs upstream of the start site (**Supplementary Figure 1b**).

**Figure 3.**
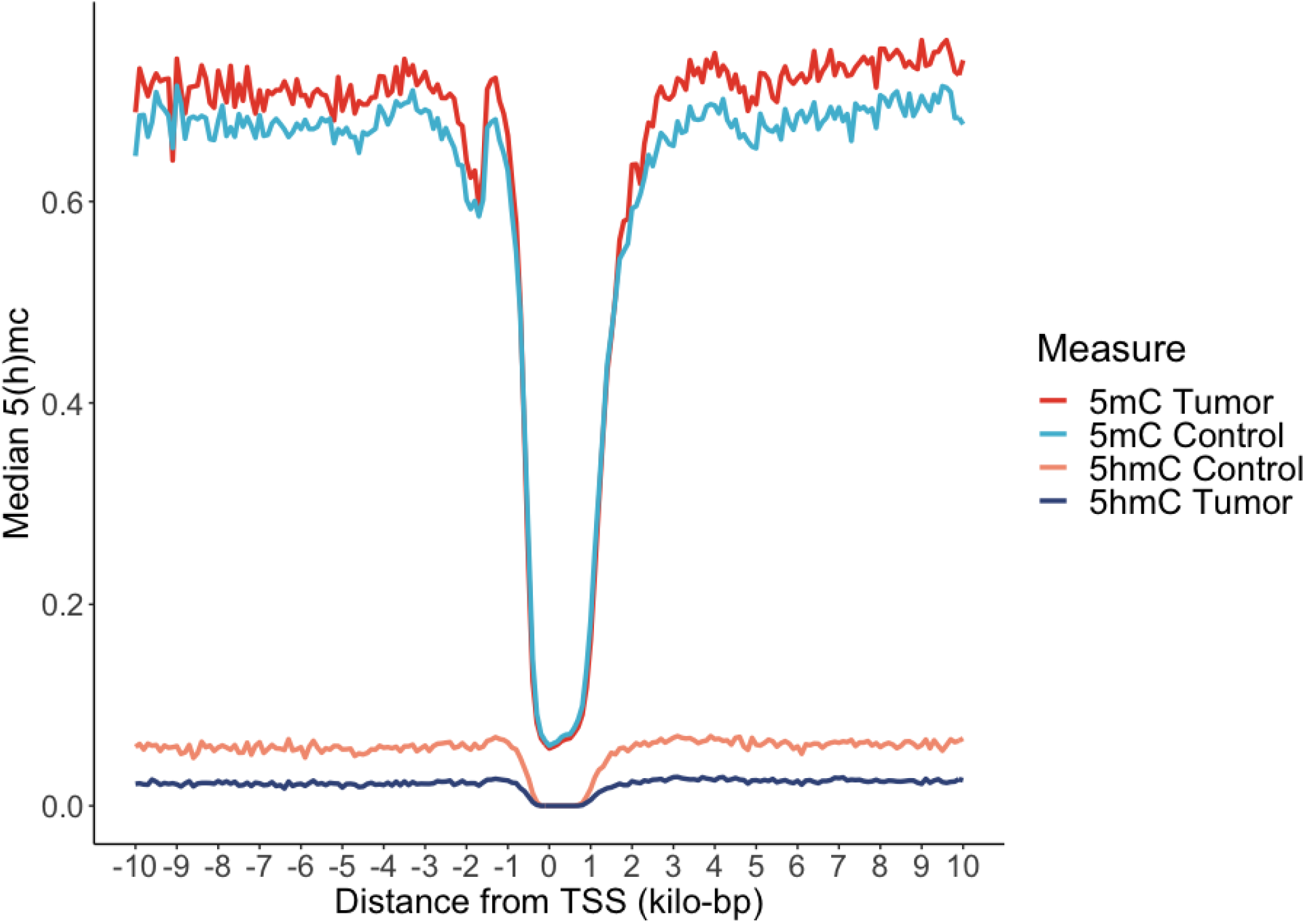
Median 5hmC and 5mC within +/- 10kilo base pairs from the nearest gene transcription start site (TSS) for both tumors (n=27) and non-tumors (n=3). We observed that there were minimal differences between tumors and non-tumors near the transcription start sites and both cytosine modifications were depleted. However, within less than 1,000 base pairs of the TSS, tumors were hypohydroxymethylated and hypermethylated as compared to controls.

We selected the top 5% of measured loci with the highest median 5hmC beta values, “high 5hmC CpGs,” for downstream analysis. This subset consisted of 37,173 loci with a median beta value greater than 0.09.

### Genomic Enrichment of 5hmC in Regulatory Regions of the Genome

We then investigated the genomic context in which CpGs with high 5hmC levels tend to cluster. Specifically, we examined their relation to CpG islands and other genomic transcriptional regulatory elements (**Figure 4**). Tumor CpG islands are significantly depleted of high 5hmC CpGs (OR = 0.03, 95% CI =0.03-0.03, P < 2.2E-16). Out of the 37,173 loci designated as high 5hmC, only 281 were identified as residing in the CpG islands. CpG depleted open sea regions are significantly enriched for high 5hmC CpGs (OR = 2.71, 95% CI = 2.64-2.78, P < 2.2E-16). In fact, 77% of high 5hmC loci were in open sea regions (**Supplementary Table 3)**. High 5hmC sites are predominantly found in genomic regions associated with transcriptional regulation such as enhancers (OR = 1.56, 95% CI = 1.49-1.64, P=7.68E-69), transcription factor binding sites (OR =1.14, 95% CI = 1.11 -1.17, P= 3.57E-20), and 5’ untranslated regions (UTR-5) (OR =1.29, 95% CI =1.25-1.33, P = 2.06E-61). Super-enhancers have been shown to be critical to cell identity, and aberrant methylation in these regions has been implicated in the development of human cancer.^42^ We accessed super-enhancer coordinates defined in multiple brain-derived cell lines from the dbSuper database and identified significant enrichment of high 5hmC including astrocyte lines (OR 1.75, 95% CI = 1.65-1.86, P = 1.94E-68) and anterior caudate lines (OR 1.93, 95% CI = 1.88-2.03, P = 6.22E-216, **Figure 4**).

**Figure 4:**
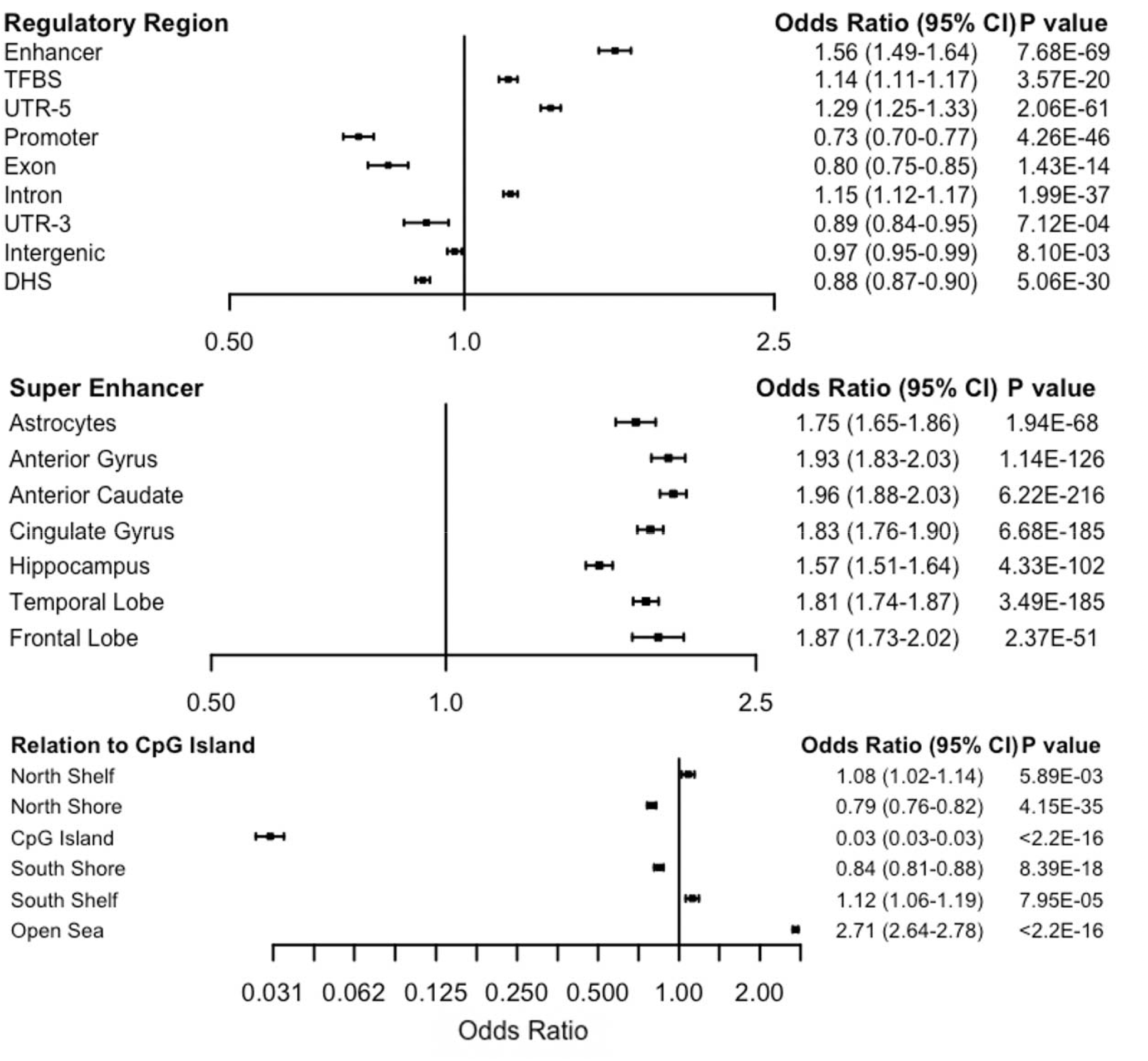
Enrichment of high 5hmC nucleotides in transcriptional regulators. These loci are modestly enriched in enhancers, transcription factor binding sites, and 5’ untranslated regions. Conversely, they are depleted in promoters. Here the top 37,173 CpGs are compared to all the sites included in the EPIC methylation array. Compared to the 850k universe, these sites are significantly depleted in CpG islands and enriched in Open Sea regions. Consistent with previous studies examining highly hydroxymethylated loci in adult glioblastomas, these CpGs are enriched in super enhancers suggesting a role in determining cellular identity.

Next, we investigated the enrichment of high 5hmC loci at the gene level. We first identified all genes with at least ten high 5hmC CpGs (n=486 genes). We defined the gene-level high 5hmC sites as a proportion of measured CpGs that were high 5hmC to account for different gene lengths, number of CpGs measured on the array, and number of high 5hmC CpGs. The range of high 5hmC sites as a proportion of those measured ranged from 1.4% to 52.6% and the mean proportion of hydroxymethylated CpGs among the 486 genes was 17%. Genes with the highest proportion of their CpGs designated as “high 5hmC” were *SHOC2* (53%), *ZRANB1* (48%), *MBNL1* (41%), *ZMIZ1-AS1* (35%), *DICER1* (35%), and *GNAQ* (35%) (**Supplementary Data 2**). This list also included, but was not exclusive to, genes pertaining to RNA interference pathways such as *DICER1* and Argonaute gene (*AGO2*)^43–45^and neurodevelopment (*GNAQ, FOXO3*).^46,47^ Using the genomic coordinates of the list of genes enriched for high 5hmC loci as the input, the Genomic Regions Enrichment of Annotations Tool (GREAT) analysis revealed biological pathways including regulation RNA interference pathways and RISC complex formation, as well as cell signaling pathways, and neuronal signaling and development. High 5hmC pathways also included those associated with craniofacial and neurodevelopmental pathologies, including autism, hyperactivity, and slanted palpebral fissures (**Supplementary Data 2**).

### Locus Specific Differentially Hydroxymethylated Regions

We tested the high 5hmC sites for differential hydroxymethylation between tumor and non-tumors (n= 37,173), adjusting for age and sex using a linear model. Tumors were hypohydroxymethylated, with 84% of the high 5hmC loci demonstrating reduced hydroxymethylation, and we observed 726 CpG loci with significantly hypohydroxymethylation in tumors (FDR < 0.1, **Figure 5a**). Submitting the 726 differentially hypohydroxymethylated regions (DHMRs) to GREAT analysis resulted in overlap with steroid response pathways (4 of the 20 gene ontology biological processes), RNA stability, as well as WNT signaling and beta-catenin binding (**Supplementary Data 3**). Furthermore, in contrast to the overall genomic context patterning of high 5hmC CpGs, tumor DHMRs were significantly enriched in CpG islands and shore regions while depleted in open sea regions (**Figure 5b**). The majority of CpGs, 63%, were found in Open Sea regions (**Supplementary Table 4**). Differentially hydroxymethylated regions (DHMRs) in CpG islands were associated with 10 genes which included *APC2, KDM2A* (an H3K36 demethylase), and GPRC5B (orphan G protein-coupled receptor critical for neurodevelopment) (**Supplementary Data 3)**.^48^

**Figure 5.**
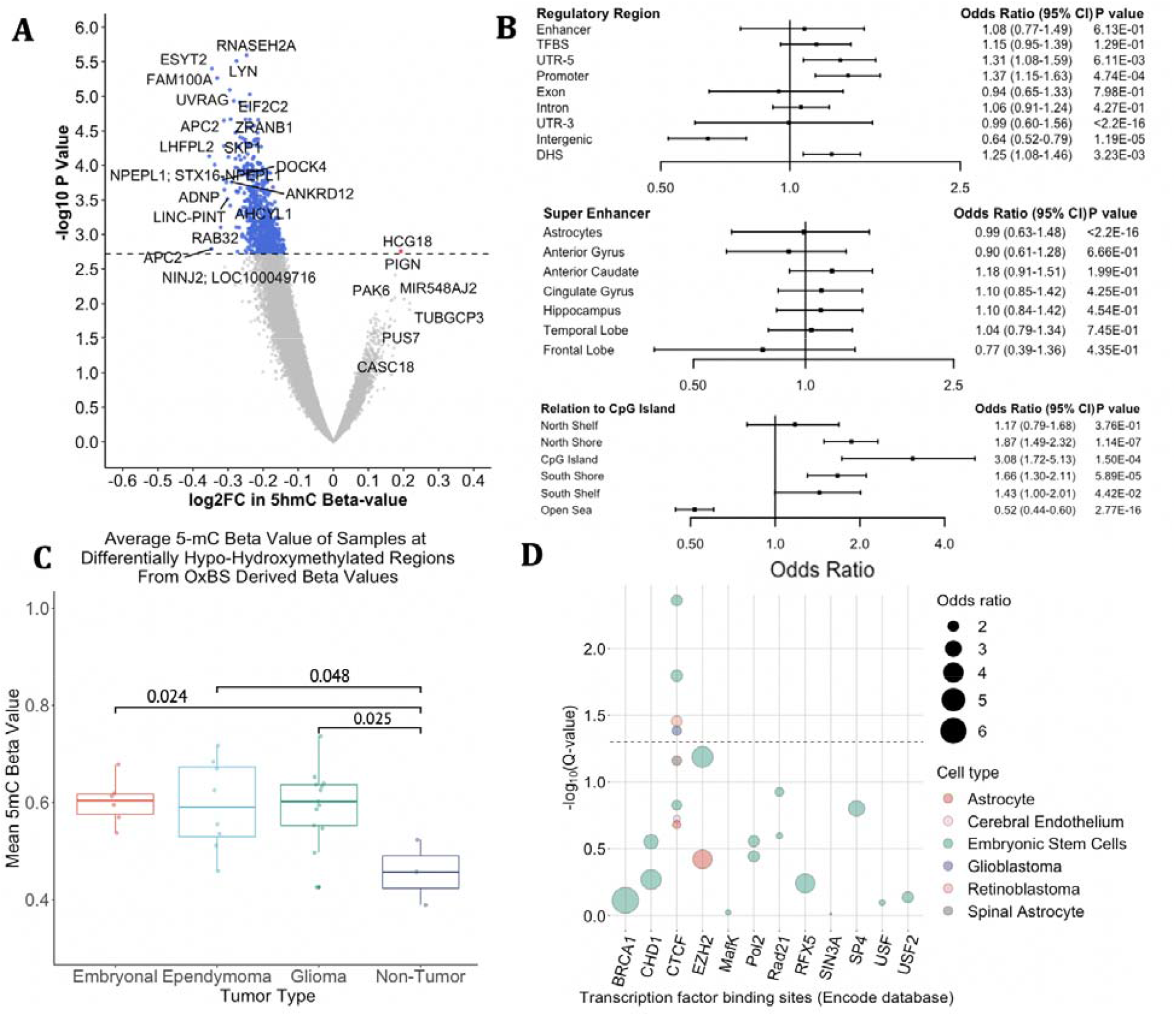
**a)** This demonstrates results of the high 5hmC CpGs (n=37,173) comparing tumor and non-tumor tissue from an age and sex adjusted linear model (limma). Volcano plot of differentially hydroxymethylated CpGs, plotting the difference of log2 fold change in beta value and the respective negative log10 of the unadjusted p-value. Blue points represent 726 differentially hypohydroxymethylated regions (DHMR) with an adjusted p-value (FDR) < 0.1. **b)** Forest plot demonstrates enrichment of differentially hydroxymethylated regions within the high 5hmC loci. These DHMRs are significantly less likely to be in intergenic regions and more likely to be in the vicinity of CpG islands. **c)** Oxidative bisulfite derived mean 5mC beta values at these DHMRs stratified by tumor type demonstrates significant hypermethylation at these loci in tumors as compared to controls. However, bisulfite only cannot distinguish between 5mC and 5hmC and demonstrated no change or hypomethylation. **d)** We previously demonstrated enrichment of high 5hmC loci in transcription factor binding sites. Here we leverage the locus overlap analysis package (LOLA) to determine the significance of overlap of hypohydroxymethylated sites with binding sites of specific transcription factors profiled by ENCODE. The top 12 enriched transcription factors discovered by LOLA analysis are represented above. TFs are plotted on the x-axis and the y axis represents the -log10 q-value (corrected for multiple hypothesis testing). The size of the bubbles represents the odds ratio and the color represents the cell line. The dotted line represents a q value of 0.05. CTCF is the only transcription factor that meets statistical significance.

To identify if particular transcription factors are associated with DHMRs, we tested for enrichment in binding sites from transcription factor experiments (ENCODE) using the Locus Overlap Analysis (LOLA) software in CNS and embryonic stem cell lines. We identified enrichment for DHMRs at CTCF binding sites, a transcription factor linked to alternative splicing by regulating RNA polymerase II and TET^49^ (**Figure 5d**). Using LOLA to interrogate sites associated with histone modifications generated by the NIH Roadmap Epigenomics Project, we found that DHMRs were significantly associated both with marks of transcriptional repression (i.e. H3K4me3) and priming (H3K4me1) (**Supplementary Figure 2**). Lastly, we investigated total 5mC levels among DHMRs. All tumor subtypes were hypermethylated at these sites as compared to non-tumors (**Figure 5c**). This pattern of hypohydroxymethylation and hypermethylation was only captured in tandem OxBS derived data and was lost in BS only derived data with the latter demonstrating a relative hypomethylation in tumors (**Supplementary Figure 3)**. In tumors, these sites also demonstrated higher levels of 5mC compared to average 5mC levels in the rest of the genome (**Supplementary Figure 4**).

### Subtype to Non-Tumor Comparisons

To further explore high 5-hmC sites (n=37,173) in tumor and non-tumor tissues, we stratified by tumor type (embryonal, ependymoma, glioma) and compared to non-tumor tissues adjusting for age and sex. Embryonal tumors had the most extensive differential hypohydroxymethylation with 12,593 loci (40%) losing 5hmC (FDR < 0.1, (**Supplementary Figure 5a**). We subset these loci to CpGs with an FDR < 0.02 (n = 1,832) and performed a GREAT analysis. The previously observed response to steroid pathways was maintained, and pathways related to mTOR regulation emerged (**Supplementary Data 4**). The extent of differential hydroxymethylation in ependymomas compared with non-tumor tissues was lower than that of embryonal tumors (n=1,630 DHMR, FDR < 0.1, **Supplementary Figure 5b**). We did not observe significant differential hydroxymethylation among gliomas compared with non-tumor samples’ DHMRs (**Supplementary Figure 5c**). We repeated GREAT analysis for ependymomas with loci with an FDR < 0.1 (n=1,630) and gliomas with an FDR < 0.2 (n= 637) (**Supplementary Data 4)**. Response to corticosteroid pathways was consistently present across all tumor types.

### 5hmC and Survival

Prior publications identified an association between decreased total 5hmC and survival in CNS tumors with mixed results.^20,40^ Here, we used a model-based clustering method, Recursively Partitioned Mixture Model (RPMM),^50^ to discern profiles of 5hmC and investigate the association of profiles with survival. Applying RPMM to the top 10,000 most variable sites in tumor samples yielded two classes of tumors, which correlated with mean 5hmC levels (p= 2.735E-05, Kruskal–Wallis rank-sum test), and we then defined the two classes as high 5hmC and low 5hmC. Class membership was not associated with tumor type, subject age, or tumor grade. The low 5hmC cluster was composed of 25% embryonal tumors, 25% ependymoma, 50% gliomas while the high 5hmC class had 18% embryonal, 36% ependymoma, 45% gliomas (**Figure 6a)**. Patients whose tumors were in the low 5hmC class had significantly poorer survival (log-rank p = 0.039, **Figure 6b**) and shorter time to disease recurrence (log-rank p=0.05, **Figure 6c**). Then, we fit two independent multivariable Cox proportional hazards models for survival and recurrence adjusting for patient sex and tumor type, for the outcomes of death and recurrence. The low 5hmC cluster was associated with an increased hazard of death (HR=6.47, 95% CI 0.79– 53.2, P = 0.08, Concordance Index =0.84, Cox proportional hazards regression) and recurrence (HR =4.83, 95% CI 0.95 – 24.6, P = 0.06, Concordance Index = 0.79) (**Supplementary Table 5**).

**Figure 6.**
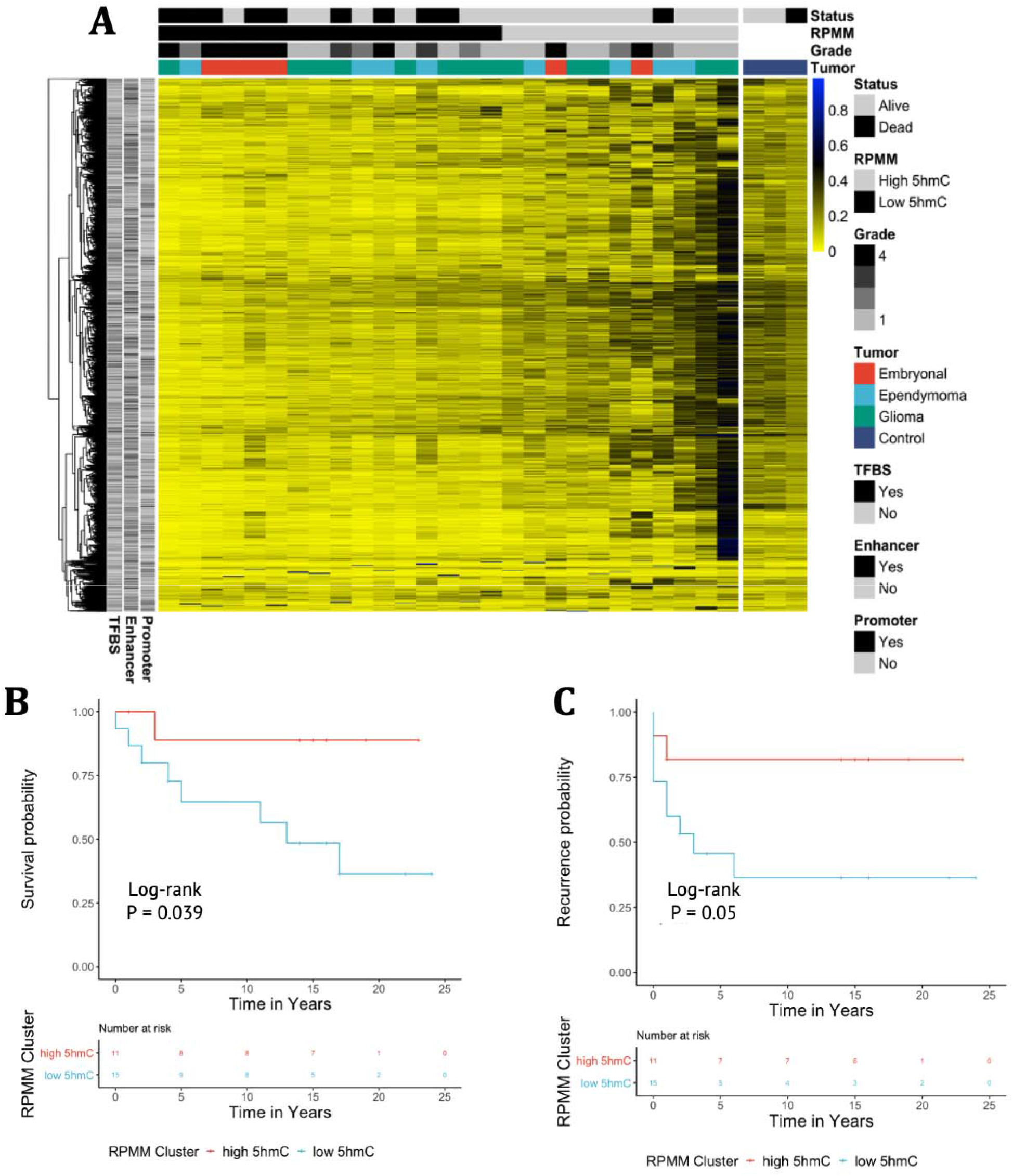
**a)** Recursively partitioned mixture model (RPMM) of tumor samples of the top 10,000 most variable CpGs in tumor samples. In the heatmap, each column represents an individual sample and each row is a CpG. Two clusters emerged that correlated with average 5hmC and were so labeled high 5hmC (gray) and low 5hmC (black). Tumors and controls are separated by a space. Patient status as alive or dead is denoted under Status and tracks with RPMM cluster as does tumor grade. Location marks the tumor as super or supratentorial. **b)** Kaplan-Meier survival plot for RPMM clusters of low 5hmC (blue) and high 5hmC (red). Association with the high 5hmC class was associated with improved survival. A multivariate cox proportional hazard model adjusting for age, sex and tumor type demonstrated an increased hazard of death for the low 5hmC RPMM cluster (HR 6.47, 95% CI 0.79-53.2, P= 0.08). **c)** Kaplan Meier plot of time to recurrence for RPMM clusters of high and low 5hmC. Trends demonstrate a longer recurrence free survival of those in the high 5hmC cluster. A multivariate cox proportional hazard model adjusting for age, sex, and tumor type demonstrated an increased hazard of recurrence for the low 5hmC cluster (HR 4.83, 95% CI 0.95 – 24.6, P=0.06).

In order to compare our results to more common methods of correlating 5hmC with survival, we constructed an index of total 5hmC by averaging beta-values of all CpGs (n= 743,461) and assigning a sample to a “High Total 5hmC” group if the mean was greater than the 50^th^ percentile (i.e., the sample mean 5hmC level was greater than 0.035). The distribution of tumor types in the low 5hmC category was 29% embryonal, 29% ependymoma, and 43% glioma; in the high 5hmC group, it was 17% embryonal, 33% ependymoma, 50% glioma. Results were consistent with the RPMM method, with the low 5hmC group having poorer survival (log-rank p=0.08, concordance index 0.75) **(Supplementary Figure 7)**. In a multivariable Cox proportional hazards model adjusted for sex and tumor type, the low total 5hmC group was associated with increased risk of recurrence (HR = 3.6, CI 95% 0.9-14.5, P = 0.07) and death (HR=3.4, CI 95% 0.69-16.5, P = 0.14).

Principle component analysis (PCA) of this subset of CpGs, revealed discernible clusters that overlapped substantially with the different tumor types (**Supplementary Figure 6a**). PCA also showed an overlap of clusters with survival (**Supplementary Figure 6b**). Using methylation beta values at these loci yielded adequate clustering by survival as well (**Supplementary Figure 6c**).

## Discussion

Childhood nervous system tumors have a relatively low incidence of mutations. Common variations tend to be related to epigenetic mechanisms such as chromatin remodeling.^51–53^ DNA methylation has been explored as a potential driver, and our results delineate the unique role of 5hmC from 5mC in CNS tumor pathology. We demonstrate that 5hmC is lost in tumors in a locus-specific pattern and that these sites are hypermethylated. This mirrors findings in adult glioblastomas, which undergo a similar exchange of 5hmC for 5mC.^30^ Oxidative bisulfite treatment enabled us to resolve this relationship, whereas bisulfite only treated samples – which measure the sum of 5hmC and 5mC – demonstrated relative hypomethylation in tumors as compared to controls. CpG islands that were specifically affected by this pattern include *APC2*, a regulator of WNT signaling pathway and a histone demethylase, *KDM2A*. Our findings reinforce the importance of a nuanced approach to the study of cytosine modifications in cancer particularly if these markers are to be used for classification.^54^

Though 5-hydroxymethylation in pediatric brain tumors has been investigated, studies to date have not been on a genome-scale at a locus resolution. An association between 5hmC and anaplasia was established using immunohistochemistry (IHC) to examine 5hmC in all WHO classifications of brain tumors.^55^ Our study adds to existing literature that 5hmC accumulates in super-enhancers and suggests a link between a loss of 5hmC and anaplasia.^55^ Recently, high 5hmC levels were associated with worse survival in pediatric posterior fossa ependymomas.^40^ However, as this study was conducted using IHC, it is unclear where in the genome this hyperhydroxymethylation was taking place. In line with previous studies, we observed that the loss of 5hmC in tumors was associated with shorter overall survival and time to recurrence.^20,29,56^ A nucleotide level analysis allowed us to identify enrichment of high 5hmC loci in genes critical to normal craniofacial and neurodevelopment further strengthening the link between these tumors and developmental neurobiology.^57^ We also found that differentially hypohydroxymethylated CpGs are enriched in molecular pathways that are frequently connected to childhood brain tumors, particularly those related to *WNT* signaling and beta-catenin binding, implicating such genes as *APC2, WNT4, WNT11, WNT5B, SHH*, and *BCL9*. 5hmC loss in *WNT* has been associated with other tumors such as melanoma and colorectal cancer.^33,37^

The tendency of 5hmC to accumulate at 5’ splicing sites in the exon-intron boundary has been suggested as a link between the epigenetic marker and alternative splicing.^58^ We found that not only did our high 5hmC sites localize to 5’ untranslated regions in concordance with previous work but that these loci were enriched in genes implicated in posttranslational regulation of gene expression such as the *DICER1, AGO2*, and *EIF2C2*. The association between 5hmC and CTCF, a methylation-sensitive transcription factor linked to alternative splicing and RNA polymerase II regulation, has also been reported in embryonal cells.^19,59^ Increased 5hmC levels have been tied to reduced binding of nucleosomes to DNA and reduced CTCF attachment.^60^ 5hmC has been shown to oscillate at 150 nucleotides, the length of nucleosome wound DNA, and 5hmC has been suggested as a linker binding CTCF to DNA.^61^

Our study is limited by the sample size of our cohort due to the rarity of such tumors. In the future, a larger multi-institutional study is warranted to narrow the confidence interval for 5hmC association with patient outcomes and increase power for differential hydroxymethylation analysis in tumor subgroups. We did not observe significant differential hydroxymethylation among gliomas compared with non-tumor samples, which is not unlikely a reflection of the heterogeneity of this tumor class. These tumors were diagnosed prior to the WHO 2016 Classification of CNS tumors^3^ with the updated molecular studies that it entailed, and it is possible that some of these tumors would be diagnosed differently if the most recent criteria were applied. However, histopathological re-review was conducted for all samples. Though potential variation in the proportion of nontumor tissue content may have contributed to more proximal clustering with non-tumor samples and higher total 5hmC levels, the observed association of 5hmC with survival and recurrence was robust to this potential variation.

Including multiple types of pediatric central nervous system tumors, both rare and common, and comparing tumor tissue to control samples enhances the genrealizability of our findings. In the future, 5hmC measures have promise for clinical applications guiding both diagnosis and treatment.

Some samples were collected following treatment, such as the glioblastoma sample taken at the time of autopsy, and this may also contribute to variation in 5hmC and 5mC levels observed. For instance, GREAT analysis of differentially hydroxymethylated regions in the tumor versus non-tumor comparison demonstrated the clustering of DHMRs in steroid response pathways. This is likely due to the treatment of most central nervous system tumors with dexamethasone in order to reduce edema and the risk of herniation. This relationship was present across all tumor subtypes. 5hmC has been associated with steroid-induced osteonecrosis and endometriosis, and our data provide further evidence of the connection between the medication and the epigenomic marker.^62,63^

To our knowledge, we describe for the first time a genome-wide cytosine-specific analysis of 5hmC in three classes of childhood CNS tumors: embryonal, glioma, and ependymoma. Super-enhancer targeting by 5hmC in all three classes of tumors was identified, and genes commonly implicated in pediatric CNS tumors were differentially hypohydroxymethylated. We demonstrated that distinguishing methylation and hydroxymethylation is critical in identifying tumor-related epigenetic changes.

## Methods

### Study Population and Samples

Pathologically confirmed fresh-frozen primary CNS tumor specimens from 27 individuals were identified; they included thirteen gliomas, eight ependymomas, and six embryonal tumors. All patients were treated at Dartmouth Hitchcock Medical Center. Samples were collected from patients who had provided consent for the use of tissues for research purposes as approved by the committee for the protection of human subjects (Institutional Review Board). Detailed information about each patient, including demographics, tumor histopathology, survival, time to recurrence, metastasis, chemotherapy and radiation regimens, was collected from the electronic medical record system. Pathologic re-review confirmed histopathologic tumor type and grade for all cases according to the 2016 WHO classification of CNS Tumors.^3^ A table with complete patient information, including survival and treatment received is available (**Supplementary Data 1**). Four fresh-frozen non-tumor brain specimens from children ranging in age from newborn to 11 years were selected as control tissues. Three tissues were from patients with epilepsy who underwent surgical resection and were used in our analyses. One tissue was excluded from analysis as it was from a non-immune hydrops fetalis subject at only 21 6/7 weeks gestational age.

### DNA Extraction and Modification

Tumor DNA was extracted from fresh frozen tumor tissue using DNeasy Blood and Tissue Kit (Qiagen) following the manufacturer’s instructions. Approximately 1-15 mg of tumor tissue was used for DNA extraction. DNA was subjected to tandem bisulfite and oxidative bisulfite (oxBS) conversion using the TruMethyl oxBs module kit (Cambridge Epigenetix - Nugen), according to the TrueMethyl protocol M01481, Revision v2.

### *TERT* Promoter and *H3F3A* sequencing

Tumor samples were sequenced for *TERT* Promoter (C2505, C280T) and *H3F3A* mutations status (K27M, G34) using PCR amplification and Sanger sequencing. ∼10 ng of genomic DNA per sample was amplified (see Appendix for the list of primers). For *H3F3A*, OneTaq Hot Start 2x Master Mix with standard buffer was used (NEB). PCR cycle parameters were as follows: denaturation at 95 °C for 30 seconds, followed by 35 cycles of 94 °C for 30s, 48 °C for 30s, 68°C for 1 min, with the final extension at 68 °C for 5 min. Amplicons were visualized in Agarose Gel each time. For *TERT* Promoter, OneTaq Hot Start 2X Master Mix with GC buffer (NEB) was used. Cycle parameters were as follows: denaturation at 94 °C 30s, followed by 35 cycles of 94°C 30s, 59°C for 30s, 68°C for 1 min, with a final extension at 68°C for 5 minutes. DNA was purified using the Qiagen PCR purification kit prior to visualization on an agarose gel. Primer sequences used can be found in **Supplementary Data 5**.

### Cytosine Modification Measures

Illumina Human Methylation EPIC Beadchips were loaded with sample DNA in a randomized fashion with equal distribution of all tumor subtypes and non-tumor samples in each chip in order to eliminate batch effects. Beadchips were read using the Illumina iScan reader, and sample intensity data (IDAT files) was generated. Normalization and background correction from each of the BS and oxBS converted samples was performed using the *Funnorm* procedure available in the R/Bioconductor package *minfi* (version 1.30.0).^64^ Before analysis, we removed CpG sites on sex chromosomes using the updated Illumina EPIC annotation file (R/Bioconductor package *IlluminaHumanMethylationEPICanno*.*ilm10b4*.*hg19*) and SNP probes identified in the EPIC array.^65^ To estimate methylated, hydroxymethylated, and unmethylated proportions of cytosine, we applied the OxyBS algorithm.^39^ Through this method, a total of 743,461 probes were left for analysis. Array data analysis was conducted in R version 3.6.1.^66^

### Statistical Analyses

We identified and subset our data to the 5% (37,173 / 743,461) most highly hydroxymethylated CpG sites (high 5hmC loci), as determined by the median, across all tumor types. We performed Fisher’s exact tests for enrichment of these 37,173 consistently hydroxymethylated CpGs in CpG islands, shores, and shelves against the 743,461 CpG universe. For gene promoter, exon, and intron regions and regulatory elements, we used a Cochran-Mantel-Haenszel test to assess enrichment stratifying by probe type. The Phantom 5 enhancer annotation (available in the Illumina EPIC annotation file) was used to map CpGs in enhancer regions. Coordinates for super-enhancers were downloaded from the dbSuper database.^67^ We examined the 486 genes in which these high 5hmC CpGs were enriched using the UCSC reference gene names in the Illumina annotation file. We selected for genes that had at least 10 CpGs represented in the EPIC array. We used the genomic coordinates of these high 5hmC CpGs in enriched genes as a query set of regions and submitted to Genomic Regions Enrichment of Annotations Tool (GREAT) analysis. We tested for enrichment against the background of the 743,173 CpGs in the EPIC array used in our analyses.

### Analysis of CpG-specific Associations

Differential hydroxymethylation status between tumor and non-tumor brain tissue at the CpG loci in our data set was determined through multivariable linear models for microarray data (limma).^68^ Models were adjusted for subject age and sex and applied to 5hmC β-values. Benjamini-Hochberg correction was used to adjust for multiple testing. We examined whether there were differences between tumors and non-tumors in 5hmC levels within the top 5% most hydroxymethylated CpGs (37,173). We applied the same model to each tumor subtype and compared them to non-tumor samples in order to determine if the results changed when stratified.

To explore if the loss of tumor hydroxymethylation in high 5hmC loci was limited to specific gene sets, we selected CpGs that were differentially hypohydroxymethylated with negative log fold change (log-FC < 0) and FDR less than 0.1 (726 CpGs) and queried the GREAT software.^69^ To test if these loci were associated with transcription factor binding sites or sites of histone modifications, we interrogated the ENCODE and Roadmaps database using the LOLA R/Bioconductor package.^70^ We tested for enrichment in CpG islands, shores and shelves using a Fisher’s exact test against the 37,173 high 5hmC loci. We used the same universe to test for enrichment of DHMRs in regulatory regions and super-enhancers using a Cochran-Mantel-Haenszel test.

### Survival Analysis

Recursively Partitioned Mixture Model (RPMM) has been used for clustering DNA methylation and hydroxymethylation data to identify classes of tumors based upon beta values.^50^ Here we applied RPMM to the top 10,000 most variable CpGs across tumors as determined by variance, and the resulting clustering solution contained two distinct clusters, defining a low and a high 5hmC cluster. A multivariable Cox proportional hazards model adjusting for age at diagnosis, sex, and tumor type was used to determine if cluster designation correlated with survival.

Although one patient in our cohort had a diagnosis of glioblastoma, the sample was collected at autopsy and no clinical information was available. Therefore, due to the lack of follow-up, we excluded this patient from our survival analysis.

## Data Availability

The pediatric tumor and non-tumor microarray data (IDAT files) were deposited at the Gene Expression Omnibus (GEO) under GSE152561 (http://www.ncbi.nlm.nih.gov/geo/). Data will be made available at publication.

## Author contributions

All authors contributed to the study conception and design. NA and LNN procured the samples and clinical data. NA and LP carried out laboratory experiments. NA, CLP, YC performed statistical analyses. All authors analyzed and interpreted the results. NA wrote the manuscript with contributions and critical revisions from all authors.

## Corresponding author

Correspondence to Brock C. Christensen.

## Competing interests

The authors declare no competing interests.

## Funding

This research was supported by a Prouty Pilot Award from the Norris Cotton Cancer Center at Dartmouth-Hitchcock Medical Center. National Institutes of Health R01CA216265, R01GM. Mr. Petersen is supported by the Burroughs-Welcome Fund: Big Data in the Life Sciences at Dartmouth. Ms. Azizgolshani is supported by the S.M. Tenney Fellowship at Dartmouth.

## Notes

### Competing Interest Statement

The authors have declared no competing interest.

### Author Declarations

Samples were collected from patients who had provided consent for the use of tissues for research purposes as approved by the committee for the protection of human subjects (Institutional Review Board) at Dartmouth Hitchcock Medical Center.

